# Longitudinal changes in brain diffusion MRI indices during and after proton beam therapy in a child with pilocytic astrocytoma: a case report

**DOI:** 10.1101/2021.02.09.21251092

**Authors:** Lisa Novello, Nivedita Agarwal, Stefano Lorentini, Sabina Vennarini, Domenico Zacà, Anna Mussano, Ofer Pasternak, Jorge Jovicich

## Abstract

**Background:** Proton beam therapy (PBT) is an effective pediatric brain tumor treatment. However, resulting microstructural changes within and around irradiated tumors are unknown. We retrospectively applied Diffusion-Tensor-Imaging (DTI) and Free-Water-Imaging (FWI) on diffusion-weighted Magnetic Resonance Imaging (dMRI) data to monitor microstructural changes during-PBT and after 8 months, in a pilocytic astrocytoma (PA) and normal-appearing white matter (NAWM).

**Methods:** We evaluated conventional MRI and dMRI-derived indices from six MRI sessions in a child with a hypothalamic PA: at baseline (t0), during-PBT (t1-t4), and after 8 months (t5). Tumor voxels were classified as “solid” or “fluid” based on FWI.

**Results:** While during PBT tumor volume remained stable, dMRI analyses identified two different response patterns: i) an increase in fluid content and diffusivity with anisotropy reductions in solid voxels at t1, followed by ii) smaller variations in fluid content but higher anisotropy in solid voxels at t2-t4. At follow-up (t5), tumor volume, its fluid content, and diffusivity in solid voxels increased. NAWM showed dose-dependent microstructural changes.

**Conclusions:** The use of dMRI and FWI showed complex dynamic microstructural changes in the irradiated mass during PBT and at follow-up, opening new avenues in our understanding of radiation-induced pathophysiologic mechanisms in tumor and surrounding tissues.

## INTRODUCTION

Proton Beam Therapy (PBT) is an effective novel approach for the treatment of pediatric pilocytic astrocytoma (PA).[1-3] However, little is known about microstructural changes that occur during PBT[4] within irradiated tumors and surrounding normal tissues.[5] Diffusion-weighted Magnetic Resonance Imaging (dMRI) data allows non-invasive monitoring of microstructural changes[6,7] by measuring water displacement. We investigated PBT-induced microstructural changes within a PA and in normal-appearing white matter (NAWM). We quantified changes using the widely used Diffusion-Tensor-Imaging[8] (DTI) analysis. In addition, we applied Free-Water-Imaging (FWI) analysis[9,10] which allowed us to separately characterize fluid voxels from solid PA based on Free-Water (FW) content to account for treatment- or disease-associated extracellular water accumulation.

## MATERIALS AND METHODS

A child (age range: 10-15 yrs) with a histologically determined PA underwent PBT six months after partial tumor resection. A ventriculoperitoneal shunt (VPS) was placed to prevent hydrocephalus. PBT was delivered in 30 daily conventional fractions of 1.8Gy Relative Biological Effectiveness (RBE) for a total target cumulative dose of 54Gy. Treatment planning details are provided in the Supplementary Materials.

### Imaging protocol

Brain structural MRI and dMRI data were acquired (1.5T Philips Ingenia) before PBT (t0), four times during a one-month-long PBT (t1, t2, t3, and t4, Table 1) and at 8-months follow-up (t5).

**Table 1:** Longitudinal quantification of brain tumor microstructural changes during and after proton therapy treatment. Each diffusion index reports the median value of the distribution within the whole tumor volume (*i.e*. including fluid voxels), the relative change of the median with respect to baseline (Δ), with its 1st (Q1) and 3rd (Q3) quartile values. Abbreviations: FA (Fractional Anisotropy), FAt (tissue FA), MD (Mean Diffusivity), FW (Free Water fraction).

|  |  | TIME POINTS (t) |  |  |  |  |  |
| --- | --- | --- | --- | --- | --- | --- | --- |
|  |  | BASELINE | PROTON THERAPY TREATMENT |  |  |  | FOLLOW-UP |
|  |  | t0 | t1 | t2 | t3 | t4 | t5 |
| **dMRI acquisition from treatment start (days)** |  | - | 9 | 17 | 24 | 30 | 269 |
| **Cumulative dose (Gy) (percentage of total dose)** |  | none | 16.2 (30%) | 30.6 (56%) | 43.2 (80%) | 54 (100%) | none |
| **FA** | median | 0.153 | 0.136 | 0.141 | 0.159 | 0.182 | 0.118 |
| | $\Delta$ | - | -11.1% | -7.8% | +3.9% | +19% | -22.9% |
|  | Q1 | 0.104 | 0.1 | 0.099 | 0.108 | 0.126 | 0.084 |
|  | Q3 | 0.214 | 0.181 | 0.193 | 0.225 | 0.255 | 0.172 |
| **FAt** | median | 0.344 | 0.321 | 0.315 | 0.337 | 0.386 | 0.367 |
| | $\Delta$ | - | -6.7% | -8.4% | -2% | +12.2% | +6.7% |
|  | Q1 | 0.247 | 0.221 | 0.208 | 0.223 | 0.269 | 0.238 |
|  | Q3 | 0.436 | 0.41 | 0.403 | 0.434 | 0.474 | 0.465 |
| **MD**<br>($\times 10^{-3}$) | median | 1.506 | 1.618 | 1.656 | 1.615 | 1.607 | 1.913 |
| | $\Delta$ | - | +7.4% | +10% | +7.2% | +6.7% | +27% |
|  | Q1 | 1.215 | 1.321 | 1.294 | 1.217 | 1.193 | 1.615 |
|  | Q3 | 1.855 | 2.191 | 2.285 | 2.211 | 2.215 | 2.249 |
| **FW** | median | 0.589 | 0.627 | 0.635 | 0.614 | 0.603 | 0.745 |
| | $\Delta$ | - | +6.5% | +7.8% | +4.2% | +2.3% | +26.5% |
|  | Q1 | 0.448 | 0.492 | 0.471 | 0.426 | 0.407 | 0.628 |
|  | Q3 | 0.726 | 0.809 | 0.841 | 0.815 | 0.811 | 0.857 |

MRI acquisitions included anatomical 3D-MPRAGE T1-weighted (T1w), 3D-FLAIR, TSE T2-weighted (T2w) and dMRI. Sequence parameters are described in Table S1. Imaging was performed for continuous monitoring of target volumes over the treatment course and to assess re-planning needs. The patient’s parents signed a written institutional consent to use MR data for research purposes.

### Patient and Public Involvement

The patient and parents were not involved in the research design nor in its questions formulation or outcome dissemination.

### Image analysis

#### Shunt-artifact mask

A VPS-induced artifact was visible (Fig. 1A, Fig. S1, S2, S3), manually masked, and excluded from all analyses.

**Fig. 1:**
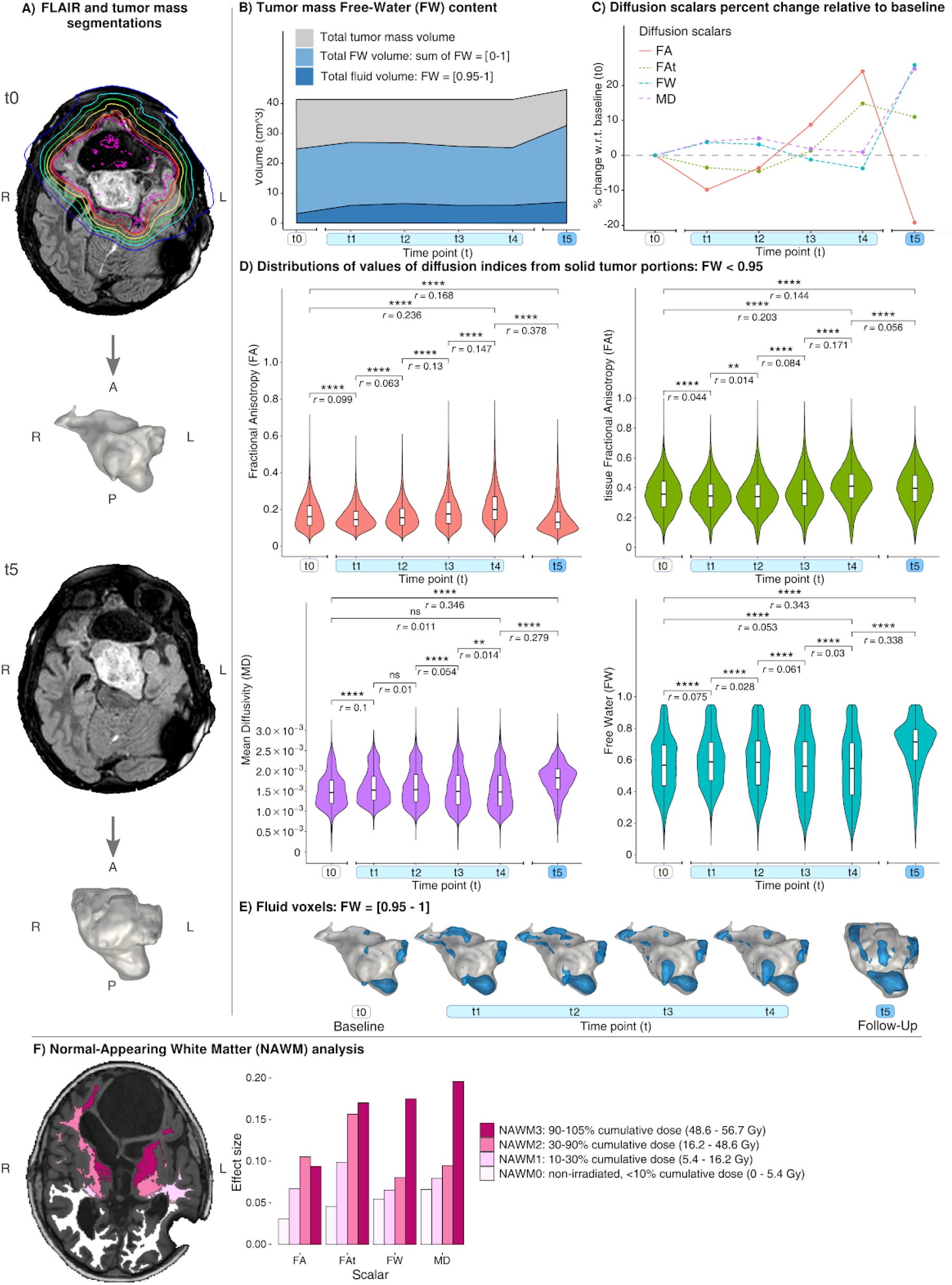
**A)** FLAIR images from baseline (t0) and 8-month post-treatment follow-up (t5). Radiation isodose curves overlaid on FLAIR at t0 (in tumor-proximal to tumor-distal order): purple: ≥100% dose; red: ≥95% dose; orange: ≥90% dose; yellow: ≥70% dose; green: ≥50% dose; cyan: ≥30% dose; blue: ≥10% dose. Below each FLAIR image the corresponding tumor mass segmentation 3D render is shown (enlarged view for better visibility). **B)** Area chart showing total tumor mass volume (grey, upper), overall FW volume (light blue, middle) and volume of voxels with FW=[0.95-1] deemed “fluid” voxels (dark blue, lower). **C)** Longitudinal percent changes with respect to baseline (t0) of median diffusion scalar values within the tumor mass (voxels with FW<0.95). **D)** Distributions of diffusion metrics in tumor mass across time points for all scalars with significance of differences between time points. Voxels corresponding to FW=(0.95-1) were excluded. The durations of each time point with respect to baseline are defined in Table 1. **E)** Mapping of FW values in dark blue in panel B) (“fluid” voxels) on tumor mass 3D render for each time point. **F)** Left: Normal-Appearing White Matter (NAWM) segmentation color-coded for irradiation dose (in tumor-proximal to tumor-distal order): magenta: NAWM3 (cumulative dose percent range: 90 - 105%); dark pink: NAWM2 (cumulative dose percent range: 30 - 90%); light pink: NAWM1 (cumulative dose percent range: 10 - 30%); white: NAWM0 (cumulative dose percent range: <10%). Right: Wilcoxon Effect size (*r*) for t0 vs. t5 comparisons for all scalars. All comparisons revealed significant (p<.0001, Bonferroni corrected) differences between baseline and follow-up for each NAWM segmented region, though with overall dose-dependent effect size values. % variation for all time points can be found on Fig. S4. Abbreviations: MD (Mean Diffusivity), FW (Free Water fraction), FA (Fractional Anisotropy), and FAt (tissue FA), ns: not significant, ** : p<.01, *** : p<.001, **** : p<.0001.

#### dMRI processing

dMRI images were denoised with Gibbs ringing (https://www.mrtrix.org/), eddy-current, motion (https://fsl.fmrib.ox.ac.uk/), and bias-field corrections. DTI[8] was used to compute Mean Diffusivity (MD) and Fractional Anisotropy (FA) maps. We used FWI[9,10] to explicitly model an extracellular free-water compartment and a tissue compartment, resulting with an extracellular FW volume fraction map (Fig. S2), and a tissue FA (FAt) map, selectively representing the FA of water molecules hindered or restricted by tissue.

#### Tumor segmentation

Tumor volumes for all timepoints were computed on manually segmented FLAIR images using 3D-Slicer (https://www.slicer.org/) by an experienced neuroradiologist (Fig. 1A). The image registration pipeline is described in the Supplementary Materials.

#### NAWM radiation dose contour volumes

NAWM was segmented from each timepoint’s T1w image into four regions based on their cumulative radiation dose percentage (Fig. 1F, and Fig. S3), as described in the Supplementary Materials.

#### dMRI longitudinal analyses

Longitudinal changes of diffusion indices from the tumor and the four NAWM regions were evaluated. Since tumor volume variations from t0 to t4 were negligible (see “conventional MRI” Results), we used the t0 tumor segmentation for during-treatment analyses. Tumor voxels with FW≥0.95 (hereafter referred to as “fluid”) were separated from those with FW<0.95 (hereafter referred to as “solid”). Fluid voxels were monitored by measuring their total volume at each timepoint. Solid voxels were monitored by investigating longitudinal differences in the DTI- and FWI-derived indices by means of Wilcoxon Rank Sum tests in R, with *p*-values Bonferroni-corrected for longitudinal comparisons and for the four diffusion indices. Longitudinal differences in DTI- and FWI-indices in all NAWM voxels were tested (t0 vs. t5) with a Wilcoxon Rank Sum test in R, and *p*-values were Bonferroni-corrected for the four diffusion metrics and the four regions investigated.

## RESULTS

PBT was well-tolerated without the need for supportive treatment. At t5 the patient presented with vomiting, headache and ideomotor slowing, which was successfully treated with corticosteroids.

### Longitudinal changes on conventional MRI

#### Baseline (t0)

T2w and FLAIR identified a hyperintense partially resected mass of 41.3 cm^3^ centered in the ventral hypothalamus surrounded by two fluid containing cysts that were hyperintense on T2w and hypointense in FLAIR, attached to the left lateral and posterior mass aspects (Fig. 1A and Fig. S1).

#### Acute PBT changes (t0 to t1)

The mass showed increased signal on T2w image. Both cysts were hyperintense on FLAIR (Fig. S1).

#### Changes during PBT (t1 to t4)

Whereas the T2w signal within the mass progressively increased, the FLAIR signal in the cysts progressively decreased (Fig. S1). Total tumor volume remained stable relative to t0: mean Dice coefficient of overlap was 0.95, mean relative volume change was 5.35% (Fig. 1B).

#### Follow-up changes (t4 to t5)

T2w signal in the mass remained high, however FLAIR signal in the cyst increased again. Total tumor volume increased to 44.7 cm^3^ (+8.1%).

### Longitudinal microstructural changes

#### Baseline (t0)

Before PBT the tumor presented with 7.6% fluid voxels (Fig. 1B).

#### Acute PBT changes (t0 to t1)

At t1, fluid voxels volume increased by 88.5% (Fig. 1B,E). In solid voxels, both DTI- and FWI-derived indices similarly showed small but significant changes: both FA and FAt decreased whereas both MD and FW increased (Fig. 1C-D).

#### Changes during PBT (t1 to t4)

Changes in fluid voxels volume between consecutive timepoints varied from −9.5% to +11% (Fig. 1B,E). From t1 to t2, FA increased but FAt decreased, MD did not vary significantly, and FW decreased. Between t2 and t4, DTI- and FWI-derived indices showed coherent trends, with small but significant increases in FA and FAt and decreases in MD and FW (Fig. 1C-D), representing opposite trends as compared to the changes between t0-t1.

#### Follow-up changes (t4 to t5)

Fluid voxels volume increased by 18.1% (Fig. 1B,E). In solid voxels, while DTI and FWI showed similar increases respectively in MD (+23.6%) and in FW (+30.7%), they showed different scales of anisotropy changes (Fig. 1C-D): DTI-FA decreased by 34.9%, and FWI-FAt decreased by 3.3%, a ten-fold smaller percent variation with respect to DTI-FA. NAWM showed longitudinal microstructural changes at t5 that were strongest in proximity to the tumor, as reported in Fig. 1F and in the Supplementary Materials (Fig. S3-S4, Table S2).

## DISCUSSION

We detected novel tissue microstructure changes in proton-irradiated PA combining conventional MRI and dMRI.

DTI and FWI revealed two distinct patterns of radiation-induced changes during PBT, the first occurring at t1 (30% dose), and the second during t2-t4 (56%-100% dose). At t1, in agreement with observed T2w changes, tumor fluid volume increased by 88%, diffusivity and extracellular water increased, and anisotropy in solid voxels decreased. These changes likely suggest cell death, as observed after 4Gy proton irradiation in vitro,[4] and damage to PA’s dense fibrillated protoplasmic astrocytes and Rosenthal fibers.[11] However, at t2-t4, we observed only modest variations of fluid voxels. Interestingly, in solid voxels, FA and FAt increased and MD and FW decreased. Such tissue microstructural changes can perhaps be explained by known complex tumor responses to PBT such as induction of cell apoptosis, astrocytic/microglial activation, PBT-induced ischemia, and cell swelling[4,12] accompanied by alterations in tumor extracellular matrix and microenvironment.[13]

At t5, total tumor volume increased by 18.1%, with increases in both fluid volume and in diffusivity within the solid voxels, reflecting signal increase in T2w images. We believe that the observed increase in tumor volume is due to increase in the fluid content and diffusivity which supports tumor pseudoprogression, known to occur in 34% of PBT PA patients.[14] In addition, a 10-fold smaller decrease in FAt relative to FA was noticed at t5. The large FA decrease is consistent with previously reported anisotropy estimation biases when not correcting for free-water,[9,10] suggestive of reduced cellularity and extracellular matrix reorganization in the long-term. However, the smaller FAt decrease, along with the large FW increase, is perhaps reflecting mostly extracellular modifications, possibly explained by edema and clearance of cellular debris.

Our follow-up dMRI findings are in agreement with data in the only other pediatric case-study that investigated PA response to PBT with dMRI over a 7-year period.[15] Despite having a single 8-month follow-up, our analyses adds novel information: i) during PBT, cumulative dose causes different dMRI-detectable sequential hyperacute processes, and ii) additional FWI analysis, compared with DTI alone, suggested different interpretation regarding the responses of the solid tumor part at follow-up, enhancing thereby our specificity and our understanding of pathophysiological mechanisms underlying proton-beam-induced injury to PA.

We acknowledge some study limitations. Between sessions, there were minor MRI variations in sequence parameters and lack of control test-retest dMRI data. NAWM subregions were obtained using dosimetric maps rather by anatomical fasciculi[7] or contralateral “control” areas given the large brain lesion and deformation both due to surgery and VPS. Future studies should consider larger patient cohorts, longitudinal cognitive evaluations, multi-shell dMRI acquisitions that improve FWI estimation, and histological validation.

In conclusion, dMRI in combination with FWI has the potential to detect subtle microstructural tissue changes in cystic tumors. A better understanding of the complex dynamic microstructural changes might help to shed light on PBT-induced damage to PA, and help clinicians to evaluate the effectiveness of PA treatment protocols. Future studies could evaluate the predictive role of tissue-specific hyperacute responses on treatment outcome.

## Supporting information

Care Checklist

## Data Availability

Data is available on reasonable request, according to restrictions imposed by patient confidentiality.

## ACKNOWLEDGMENTS

We are grateful to the patient and to his family for their participation and consent to perform analysis on data acquired. Grant support: NIH P41EB015902.

## CONTRIBUTIONS

NA, DZ, JJ conceived the study and implemented the MRI acquisition protocol. SL, SV and AM conducted the recruitment, treatment planning and treatment. NA collected the MRI data. LN performed the whole data analyses using software developed by OP. LN, NA and JJ drafted the initial manuscript. All authors contributed to the data interpretation, revision and editing of the manuscript.

## REFERENCES

1 Levin WP, Kooy H, Loeffler JS, et al. Proton beam therapy. Br J Cancer 2005;93:849–854.

2 Amichetti, M. The actual interest in radiotherapy for the utilization of proton beam, highlighting physics basis, technology and common clinical indications. Journal of Tumor 2016;4:378–85.

3 Carbonara R, Di Rito A, Monti A, et al. Proton versus Photon Radiotherapy for Pediatric Central Nervous System Malignancies: A Systematic Review and Meta-Analysis of Dosimetric Comparison Studies. Journal of Oncology 2019.

4 Wang L, Han S, Zhu J, et al. Proton versus photon radiation–induced cell death in head and neck cancer cells. Head Neck 2019;41:46–55.

5 Kralik SF, Ho CY, Finke W, et al. Radiation necrosis in pediatric patients with brain tumors treated with proton radiotherapy. AJNR Am J Neuroradiol 2015;36:1572–78.

6 Chenevert TL, Stegman LD, Taylor JMG, et al. Diffusion magnetic resonance imaging: an early surrogate marker of therapeutic efficacy in brain tumors. J Natl Cancer Inst 2000;92:2029–36.

7 Connor M, Karunamuni R, McDonald C, et al. Regional susceptibility to dose-dependent white matter damage after brain radiotherapy. Radiother Oncol 2017;123:209–17.

8 Basser PJ, Mattiello J, LeBihan D. MR diffusion tensor spectroscopy and imaging. Biophys J 1994;66:259–67.

9 Pasternak O, Sochen N, Gur Y, et al. Free water elimination and mapping from diffusion MRI. Magn Reson Med 2009;62:717–30.

10 Albi A, Pasternak O, Minati L, et al. Free water elimination improves test–retest reproducibility of diffusion tensor imaging indices in the brain: a longitudinal multisite study of healthy elderly subjects. Hum Brain Mapp 2017;38:12–26.

11 Collins VP, Jones DT, Giannini C. Pilocytic astrocytoma: pathology, molecular mechanisms and markers. Acta Neuropathol 2015;129:775–88.

12 Srinivas US, Tan BWQ, Vellayappan BA, et al. ROS and the DNA damage response in cancer. Redox Biol 2019;25:101084.

13 Krisnawan VE, Stanley JA, Schwarz JK, et al. Tumor Microenvironment as a Regulator of Radiation Therapy: New Insights into Stromal-Mediated Radioresistance. Cancers (Basel) 2020;12:2916.

14 Indelicato DJ, Rotondo RL, Uezono H, et al. Outcomes following proton therapy for pediatric low-grade glioma. Int J Radiat Oncol Biol Phys 2019;104:149–56.

15 Hou P, Zhu KH, Park PC, et al. Proton Therapy for Juvenile Pilocytic Astrocytoma: Quantifying Treatment Responses by Magnetic Resonance Diffusion Tensor Imaging. Int J Part Ther 2016;3:414–20.

