## Supplementary Materials for "Longitudinal changes in brain diffusion MRI indices during and after proton beam therapy in a child with pilocytic astrocytoma: a case report"

**ABSTRACT**

**Background**

Proton beam therapy (PBT) is an effective pediatric brain tumor treatment. However, resulting microstructural changes within and around irradiated tumors are unknown. We retrospectively applied Diffusion-Tensor-Imaging (DTI) and Free-Water-Imaging (FWI) on diffusion-weighted Magnetic Resonance Imaging (dMRI) data to monitor microstructural changes during-PBT and after 8 months, in a pilocytic astrocytoma (PA) and normal-appearing white matter (NAWM).

**Methods**

We evaluated conventional MRI and dMRI-derived indices from six MRI sessions in a child with a hypothalamic PA: at baseline (t0), during-PBT (t1-t4), and after 8 months (t5). Tumor voxels were classified as "solid" or "fluid" based on FWI.

**Results**

While during PBT tumor volume remained stable, dMRI analyses identified two different response patterns: i) an increase in fluid content and diffusivity with anisotropy reductions in solid voxels at t1, followed by ii) smaller variations in fluid content but higher anisotropy in solid voxels at t2-t4. At follow-up (t5), tumor volume, its fluid content, and diffusivity in solid voxels increased. NAWM showed dose-dependent microstructural changes.

**Conclusions**

The use of dMRI and FWI showed complex dynamic microstructural changes in the irradiated mass during PBT and at follow-up, opening new avenues in our understanding of radiation-induced pathophysiologic mechanisms in tumor and surrounding tissues.

### SUPPLEMENTARY MATERIALS

#### Proton therapy treatment

The treatment plan was created in Raystion v6 (Raysearch, Stockholm) treatment planning system (TPS) and optimized according to a single field optimization technique using three non-coplanar fields (i.e.: Gantry 325° and couch 90°, Gantry 290° and couch 30°, Gantry 90° and couch 330°). Dose was calculated with a Monte Carlo dose engine over a dose grid having spatial resolution of 1.5mm. A 3 mm margin for the creation of the planning target volume (PTV) based on the clinical target volume (CTV) was used. The plan was designed to fulfill both dose prescription to target and constraints to organs at risk located nearby the target (Fig. 1A). A PTV coverage of V98=97% (meaning the 98% of the prescribed dose covering the 97% of the PTV) was achieved in the nominal plan, while trying to maximize sparing of organs at risk: average dose to temporal lobe was 7.6Gy and 19.6Gy respectively for right and left lobe, brainstem maximum dose was 53.2Gy and average dose to left and right hippocampus was respectively 30Gy and 9.8Gy.

#### Image co-registrations for longitudinal alignment

A VPS-artifact mask was manually segmented to exclude the artifact region in registration processes (ANTs, Advanced Normalization Tools, https://github.com/ANTsX/ANTs). Each timepoint T1w was bias-field corrected and linearly registered to baseline (t0) T1w. A brain mask derived from a computed tomography acquired at baseline was registered to t0's T1w, and a non-linear registration estimated between each timepoint b0-dMRI and T1w was applied to all diffusion scalar maps, which were resized to 1 mm-isotropic, and the resulting registered images were visually inspected. Bias-field corrected FLAIR (t0-t5) were linearly registered to the first time point T1w. This allowed us to have all longitudinal data aligned: tumor segmentations, diffusion scalar maps, T1w, and FLAIR images.

##### Normal-Appearing White Matter (NAWM) radiation dose contour volumes

T1w images were segmented using FSL's FAST (https://fsl.fmrib.ox.ac.uk/fsl), and the resulting NAWM segmentations were spatially divided based on radiation dose according to PBT contours. Four NAWM volumes were defined (Fig. 1F, Fig. S3) by % of cumulative dose: 0-10% (non-irradiated volume, NAWM0), 10-30% (NAWM1), 30-90% (NAWM2), 90-105% (NAWM3 - Fig. 1F, left).

##### Microstructural changes of NAWM at follow-up

During treatment, we observed non-monotonic changes of diffusion scalars for all NAWM areas (Fig. S3). At t5, diffusion scalars changed significantly ($p<.0001$) relative to baseline, showing overall dose-dependent effect size values, effects increasing with tumor proximity (Fig. 1F, percent changes are reported in Table S3).

##### Prior presentations of this work

Preliminary results from this work were presented as posters presentations at the International Society for Magnetic Resonance in Medicine (ISMRM) 2019 meeting in Montréal (Canada), at the ISMRM Italian chapter 2019 meeting in Milan (Italy), and at the Organization for Human Brain Mapping (OHBM) 2019 meeting in Rome (Italy).

**Table S1**: MRI protocol information. In bold, parameters changed w.r.t. baseline (t0). Abbreviations: FLAIR: Fluid Attenuated Inversion Recovery; dMRI: diffusion-weighted Magnetic Resonance Imaging; TSE: Turbo Spin Echo; TE: Echo Time; TR: Repetition Time; t1-t5 are defined in Table 1.

| Sequence | Time points (t) | TE (ms) | TR (ms) | TI (ms) | Resolution (mm) |
|---|---|---|---|---|---|
| 3D FLAIR | t0, t3-t5 | 357 | 4800 | 1660 | 0.98 x 0.98 x 0.6 |
| | t1, t2 | **351** | 4800 | 1660 | 0.98 x 0.98 x 0.6 |

| Sequence | Time points (t) | TE (ms) | TR (ms) | Flip Angle | Resolution (mm) |
|---|---|---|---|---|---|
| 3D T1w | t0, t3, t4 | 4.5 | 25 | 25° | 0.45 x 0.45 x 0.8 |
| | t1, t2 | **3.2** | **7** | **8°** | **0.47 x 0.47 x 1** |
| | t5 | **3.3** | **7** | **8°** | **1 isotropic** |
| 2D T2w TSE | t0, t5 | 100 | 6059 | 90° | 0.34 x 0.34 x 5 |
| | t1, t2 | 100 | **6463** | 90° | 0.34 x 0.34 x 5 |
| | t3, t4 | 100 | **6665** | 90° | 0.34 x 0.34 x 5 |

| Sequence | Time points (t) | TE (ms) | TR (ms) | Flip Angle | Resolution ($mm^3$) | b-value ($s/mm^2$) | Number of b=0 volumes | Number of gradient directions |
|---|---|---|---|---|---|---|---|---|
| dMRI | t0, t1 | 103 | 10117 | 90° | 2 isotropic | 800 | 1 | 32 |
| | t2 | **101** | **10029** | 90° | 2 isotropic | 800 | 1 | 32 |
| | t3 | 103 | **11735** | 90° | 2 isotropic | 800 | 1 | 32 |
| | t4 | 103 | **11001** | 90° | **1.75 x 1.75 x 2** | 800 | 1 | 32 |
| | t5 | 103 | **10556** | 90° | 2 isotropic | 800 | 1 | 32 |

**Figure S1**: Conventional structural MRI showing tumor changes with different images (FLAIR, T1w, and T2w) for all time points. Acquisition parameters are listed in Table S1. R: Right; L: Left. Two cysts are visible on tumor left lateral and posterior aspects. At baseline (t0) the lateral cyst presented CSF-like signal intensity. At t1, FLAIR revealed an increase in signal intensity within the cysts. During the following part of the treatment (t2-t4), the fluid within the lateral cyst returned to CSF-like FLAIR signal intensity. At follow-up (t5) FLAIR signal intensity within the lateral cyst returned to that observed in t1. The initial rise (t1) in the FLAIR signal within the cysts might reflect an increase in proteinaceous material released by cells lining the cysts due to radiation injury.

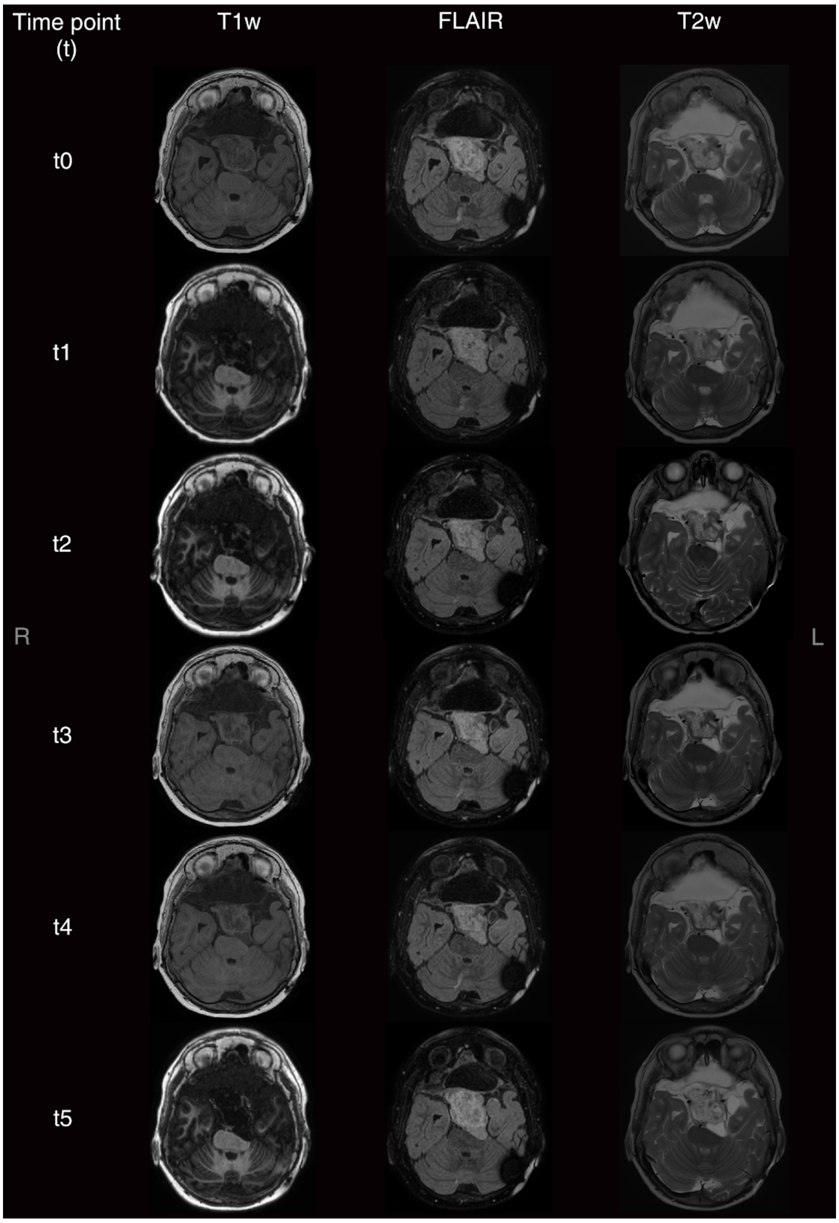

**Figure S2**: Diffusion scalar maps at baseline derived from the Tensor (left, Fractional Anisotropy and Mean Diffusivity) and the bi-tensor (right, Tissue Fractional Anisotropy, Free-Water) models.

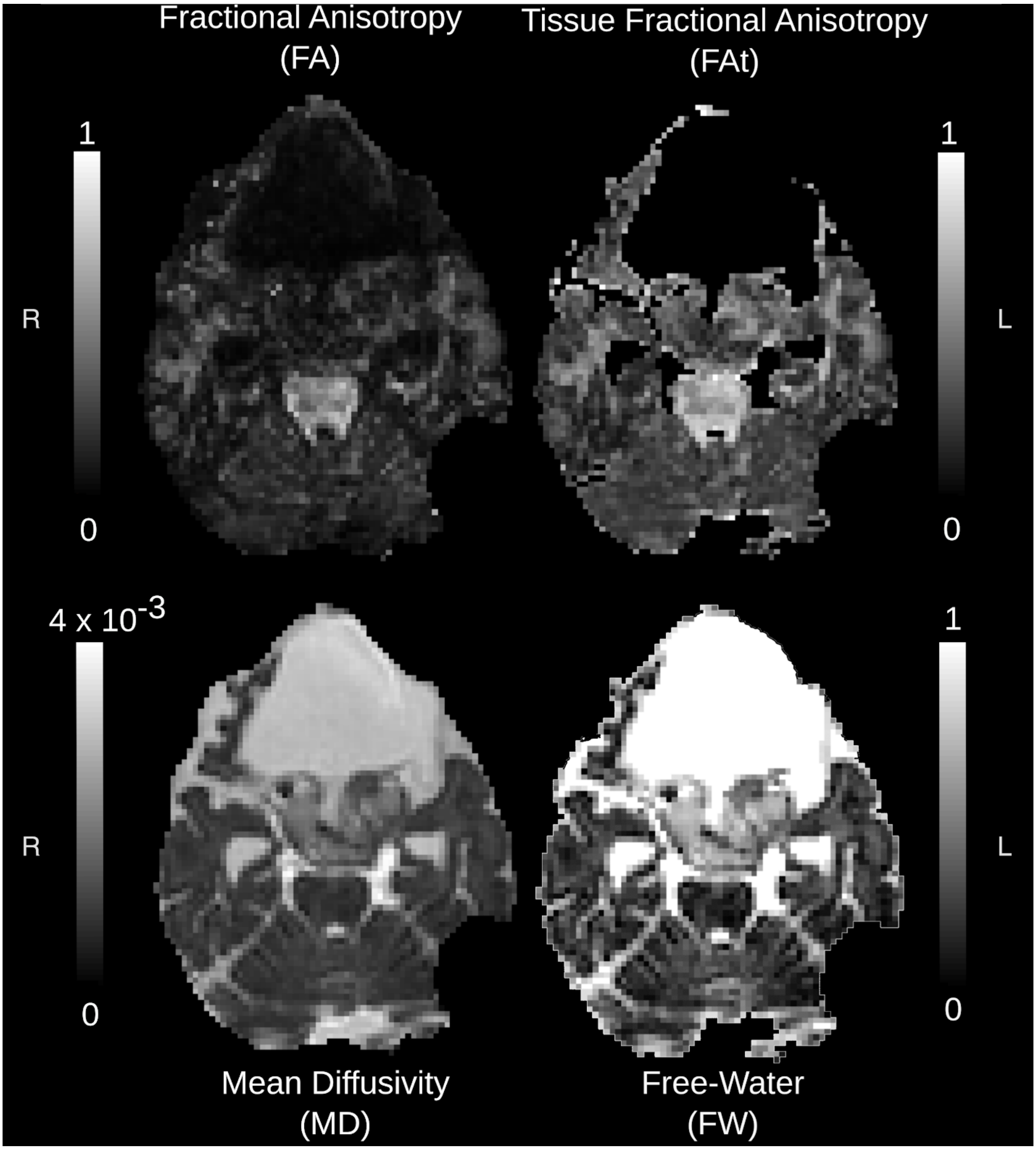

**Figure S3**: Mean Diffusivity (MD) map at t5 (8-month follow-up) with Normal-Appearing White Matter (NAWM) segmentations and tumor mass segmentation overlaid. NAWM regions (NAWM0, …, NAWM3) correspond to different cumulative radiation dose volumes. The procedure for their subdivision is described in the "*Normal-Appearing White Matter (NAWM) radiation dose contour volumes*" paragraph on the Supplementary Materials, and can be also seen in Figure 1F.

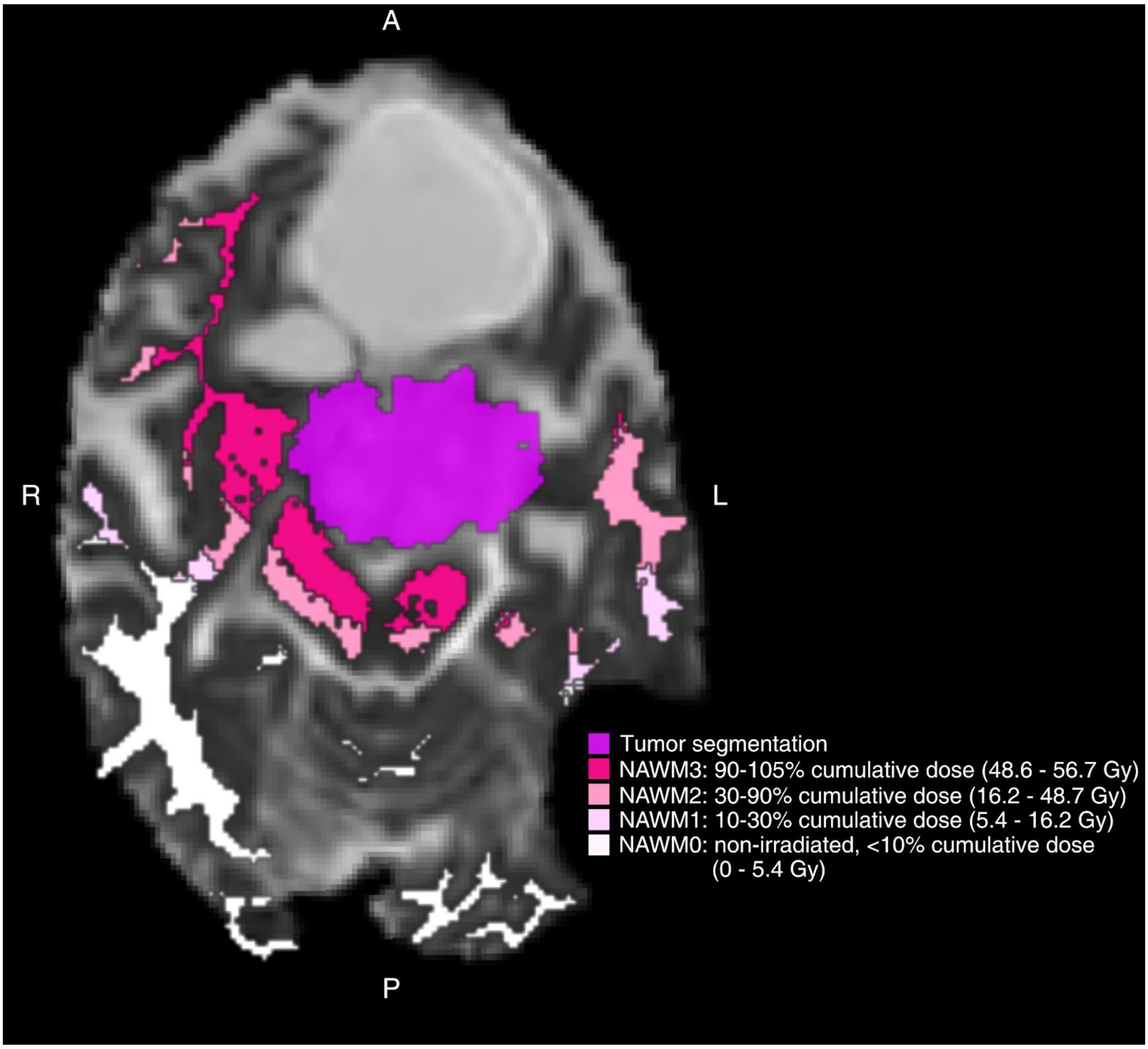

**Figure S4**: Relative changes (%) of Normal-Appearing White Matter (NAWM) diffusion metrics with respect to baseline (t0) for each diffusion scalar across time points (t1-t5, Table 1). NAWM regions (NAWM0, …, NAWM3) correspond to different cumulative radiation dose volumes and can be seen in Figure 1F. Abbreviations: FA (Fractional Anisotropy), FAt (tissue FA), MD (Mean Diffusivity), and FW (Free-Water fraction).

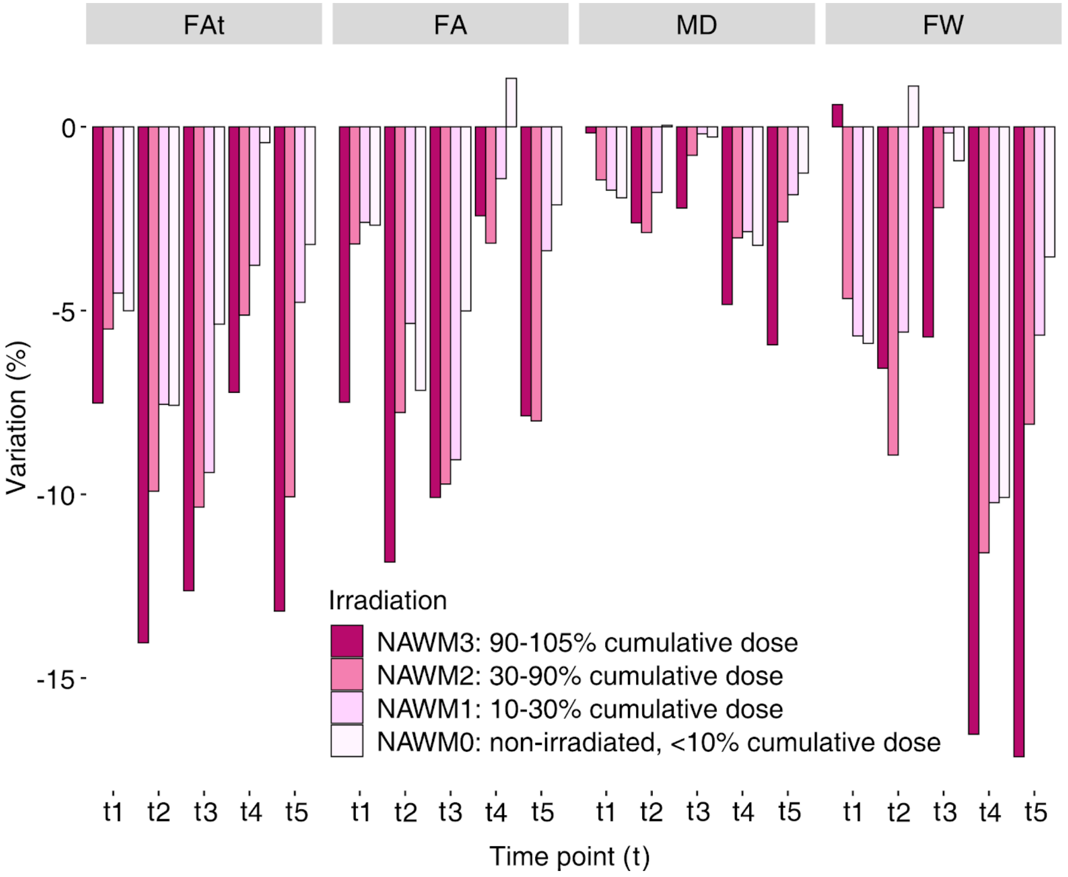

**Table S2**: Median Normal-Appearing White Matter (NAWM) diffusion metric values at baseline (t0) and relative changes (%) with respect to baseline for each diffusion scalar across time points (t0-t5, Table 1). NAWM regions (NAWM0, …, NAWM3) correspond to different cumulative radiation dose volumes (Figure 1F). Abbreviations: dMRI (diffusion MRI), FAt (tissue Fractional Anisotropy), FA (Fractional Anisotropy), MD (Mean Diffusivity), FW (Free-Water fraction).

| **dMRI metrics** | **NAWM regions** | **TIME POINTS (T)** | | | | | |
|---|---|---|---|---|---|---|---|
| | | **Baseline** | **TREATMENT** [1] (%) | | | | **FOLLOW-UP** [1] (%) |
| | | **t0** | **t1** | **t2** | **t3** | **t4** | **t5** |
| **FAt** | 0 | 0.392 | -5.01 | -7.58 | -5.37 | -0.43 | -3.20 |
| | 1 | 0.471 | -4.53 | -7.55 | -9.41 | -3.77 | -4.78 |
| | 2 | 0.507 | -5.50 | -9.92 | -10.35 | -5.13 | -10.07 |
| | 3 | 0.521 | -7.52 | -14.04 | -12.62 | -7.22 | -13.17 |
| **FA** | 0 | 0.311 | -2.67 | -7.17 | -5.01 | +1.32 | -2.12 |
| | 1 | 0.373 | -2.60 | -5.35 | -9.06 | -1.41 | -3.37 |
| | 2 | 0.395 | -3.18 | -7.77 | -9.72 | -3.16 | -8.00 |
| | 3 | 0.391 | -7.49 | -11.84 | -10.09 | -2.42 | -7.87 |
| **MD** | 0 | $0.825 *10^{-3}$ | -1.93 | +0.04 | -0.28 | -3.22 | -1.26 |
| | 1 | $0.814 *10^{-3}$ | -1.72 | -1.78 | -0.19 | -2.85 | -1.85 |
| | 2 | $0.818 *10^{-3}$ | -1.44 | -2.88 | -0.78 | -3.02 | -2.59 |
| | 3 | $0.839 *10^{-3}$ | -0.17 | -2.61 | -2.21 | -4.83 | -5.93 |
| **FW** | 0 | 0.19 | -5.89 | 1.11 | -0.92 | -10.09 | -3.54 |
| | 1 | 0.178 | -5.69 | -5.58 | -0.17 | -10.23 | -5.67 |
| | 2 | 0.179 | -4.67 | -8.93 | -2.20 | -11.59 | -8.09 |
| | 3 | 0.192 | 0.60 | -6.57 | -5.72 | -16.53 | -17.14 |

1: Percent changes relative to baseline
