## Supplementary material for "Longitudinal changes in brain diffusion MRI indices during and after proton beam therapy in a child with pilocytic astrocytoma: a case report": Care Checklist

CARE case report guidelines

### CARE Checklist of information to include when writing a case report

(cc) BY-NC-ND

| Topic | Item | Checklist item description | Reported on Line |
|---|---|---|---|
| **Title** | 1 | The diagnosis or intervention of primary focus followed by the words "case report" | 1 - 2 |
| **Key Words** | 2 | 2 to 5 key words that identify diagnoses or interventions in this case report, including "case report" | 18 |
| **Abstract (no references)** | 3a | Introduction: What is unique about this case and what does it add to the scientific literature? | 22 - 25 |
| | 3b | Main symptoms and/or important clinical findings | 29 |
| | 3c | The main diagnoses, therapeutic interventions, and outcomes | 29, 32 - 36 |
| | 3d | Conclusion—What is the main "take-away" lesson(s) from this case? | 38 - 40 |
| **Introduction** | 4 | One or two paragraphs summarizing why this case is unique (**may include references**) | 43 - 46, 49 - 52 |
| **Patient Information** | 5a | De-identified patient specific information. | 54 - 56 |
| | 5b | Primary concerns and symptoms of the patient. | 54 |
| | 5c | Medical, family, and psycho-social history including relevant genetic information | 54 - 55 |
| | 5d | Relevant past interventions with outcomes | 54 - 55 |
| **Clinical Findings** | 6 | Describe significant physical examination (PE) and important clinical findings. | 108 |
| **Timeline** | 7 | Historical and current information from this episode of care organized as a timeline | 108 - 127 |
| **Diagnostic Assessment** | 8a | Diagnostic testing (such as PE, laboratory testing, imaging, surveys). | 60 - 62, 68 - 72 |
| | 8b | Diagnostic challenges (such as access to testing, financial, or cultural) | 78 - 79 |
| | 8c | Diagnosis (including other diagnoses considered) | 54 |
| | 8d | Prognosis (such as staging in oncology) where applicable | - |
| **Therapeutic Intervention** | 9a | Types of therapeutic intervention (such as pharmacologic, surgical, preventive, self-care) | 56 - 58 |
| | 9b | Administration of therapeutic intervention (such as dosage, strength, duration) | 56 - 57 |
| | 9c | Changes in therapeutic intervention (with rationale) | - |
| **Follow-up and Outcomes** | 10a | Clinician and patient-assessed outcomes (if available) | 112 - 147 |
| | 10b | Important follow-up diagnostic and other test results | 126 - 127, 142 - 147 |
| | 10c | Intervention adherence and tolerability (How was this assessed?) | 108 |
| | 10d | Adverse and unanticipated events | 108 - 110 |
| **Discussion** | 11a | A scientific discussion of the strengths AND limitations associated with this case report | 172 - 191 |
| | 11b | Discussion of the relevant medical literature **with references**. | 172 - 179 |
| | 11c | The scientific rationale for any conclusions (including assessment of possible causes) | 151 - 171 |
| | 11d | The primary "take-away" lessons of this case report (without references) in a one paragraph conclusion | 187 - 191 |
| **Patient Perspective** | 12 | The patient should share their perspective in one to two paragraphs on the treatment(s) they received | 73 - 75 |
| **Informed Consent** | 13 | Did the patient give informed consent? Please provide if requested | **Yes** ☒ **No** ☐ |
